# Quantifying infectious disease epidemic risks: A practical approach for seasonal pathogens

**DOI:** 10.1101/2024.07.30.24311220

**Authors:** AR Kaye, G Guzzetta, MJ Tildesley, RN Thompson

## Abstract

For many infectious diseases, the risk of outbreaks varies seasonally. If a pathogen is usually absent from a host population, a key public health policy question is whether the pathogen’s arrival will initiate local transmission, which depends on the season in which arrival occurs. This question can be addressed by estimating the “probability of a major outbreak” (the probability that introduced cases will initiate sustained local transmission). A standard approach for inferring this probability exists for seasonal pathogens (involving calculating the Case Epidemic Risk; CER) based on the mathematical theory of branching processes. Under that theory, the probability of pathogen extinction is estimated, neglecting depletion of susceptible individuals. The CER is then one minus the extinction probability. However, as we show, if transmission cannot occur for long periods of the year (e.g., over winter or over summer), the pathogen will inevitably go extinct, leading to a CER of zero even if seasonal outbreaks can occur. This renders the CER uninformative in those scenarios. We therefore devise an alternative approach for inferring outbreak risks for seasonal pathogens (involving calculating the Threshold Epidemic Risk; TER). Estimation of the TER involves calculating the probability that introduced cases will initiate a local outbreak in which a threshold number of infections is exceeded before outbreak extinction. For simple seasonal epidemic models, such as the stochastic Susceptible-Infectious-Removed model, the TER can be calculated numerically (without model simulations). For more complex models, such as stochastic host-vector models, the TER can be estimated using model simulations. We demonstrate the application of our approach by considering Chikungunya virus in northern Italy as a case study. In that context, transmission is most likely in summer, when environmental conditions promote vector abundance. We show that the TER provides more useful assessments of outbreak risks than the CER, enabling practically relevant risk quantification for seasonal pathogens.

**Author Summary:** Invasive pathogens pose a challenge to human health, particularly as outbreak risks for some infectious diseases are being exacerbated by climate change. For example, the occurrence of seasonal vector-borne disease outbreaks in mainland Europe is increasing, even though pathogens like the Chikungunya and dengue viruses are not normally present there. In this changing landscape, assessing the risk posed by invasive pathogens requires computational methods for estimating the probability that introduced cases will lead to a local outbreak, as opposed to the first few cases fading out without causing a local outbreak. In this article, we therefore provide a computational framework for estimating the risk that introduced cases will lead to a local outbreak in which a pre-specified, context specific threshold number of cases is exceeded (we term this risk the “Threshold Epidemic Risk”, or TER). Since even small seasonal outbreaks can have negative impacts on local populations, we demonstrate that calculation of the TER provides more appropriate estimates of local outbreak risks than those inferred using standard methods. Going forwards, our computational modelling framework can be used to assess outbreak risks for a wide range of seasonal diseases.

## 1. Introduction

Even if a pathogen is not commonly present in a host population, there remains a risk that imported cases will lead to local transmission [1–5]. In southern Europe, for example, vector-borne diseases such as dengue and chikungunya are not endemic, yet outbreaks occur due to pathogen importation followed by autochthonous (i.e., local) transmission [6–8]. The risk that imported cases will lead to a substantial local outbreak, as opposed to sporadic onwards transmissions occurring, varies seasonally. This is because factors such as host behaviour, pathogen survivability and vector ecological dynamics change during the year, and are affected by weather variables such as temperature, rainfall and humidity [9–12]. It is useful to identify times of year at which outbreaks are most likely, and to provide quantitative estimates of temporally varying outbreak risks, to inform vector or pathogen surveillance and control interventions.

Previous work on the topic of inferring the risk that introduced cases will initiate sustained local transmission has focussed on estimating the so-called “probability of a major outbreak”, based on the number of imported cases and the transmissibility of the pathogen. This probability can be inferred both for pathogens that are transmitted directly between hosts [13–26] and those that are spread via vectors [27–30].

Furthermore, the probability of a major outbreak has been calculated in systems in which transmission parameter values are assumed to be constant [8,30–33] and those in which temporal variations in transmission are accounted for [29,34–41]. Estimates of the probability of a major outbreak have been generated using approximations of a wide range of epidemiological models, including SIS, SIR and SEIR models [30,31], spatial models [22,23,27], models with host demography [25,26,42] and models that relax the standard assumption that epidemiological time periods are drawn from exponential distributions [24,43]. In addition, calculations of the probability of a major outbreak have been undertaken for a wide variety of diseases, including COVID-19 [21,32], Ebola [31,43] and dengue [8,44].

In all these different settings, the probability of a major outbreak is typically derived by assuming that infections are generated according to a branching process [45], neglecting depletion of susceptible individuals (i.e., assuming that there is a constant supply of susceptible hosts available for each infected individual to infect). When transmission parameter values do not vary temporally, under this assumption a pathogen either goes extinct following its introduction or the number of infections grows unboundedly. The probability of a major outbreak calculated in this way corresponds to the probability that the second of these scenarios arises (i.e., that infinitely many infections occur in the branching process model). Generally, this is appropriate, and estimates of the probability of a major outbreak match the proportion of simulations of stochastic compartmental models (that account for depletion of susceptible individuals) in which “large” outbreaks occur, at least when parameters take constant values and *R*_0_ is sufficiently larger than one [29,30]. However, the use of branching process theory to estimate outbreak risks can be problematic when transmission is seasonal.

Specifically, when transmission can only occur during some periods of the year, the pathogen will inevitably go extinct in seasons when environmental conditions are unsuitable for transmission. Consequently, even with a constant supply of susceptible individuals for infected hosts to infect, the number of infections will not grow indefinitely. As a result, standard analytic estimates of the probability of a major outbreak (here called the Case Epidemic Risk, or CER, following the use of this terminology previously for pathogens for which transmission varies temporally [29]) are vanishingly small. Since pathogen extinction will almost certainly occur, a more practically relevant question is how many infections will there be before extinction? If a substantial number of infections arises prior to pathogen extinction, we contend that an outbreak should still be classified as “major”.

Here, we therefore provide a new metric for calculating the probability of a major outbreak for seasonal pathogens. Specifically, we calculate the probability that, following the introduction of a pathogen to a host population, a pre-specified, context dependent threshold number of total infections is exceeded. We refer to this metric as the Threshold Epidemic Risk (TER). This metric can be calculated using stochastic compartmental transmission models that account for both seasonality and depletion of susceptible individuals, and throughout this article we compare calculations of the TER to analogous values of the CER. A schematic is shown in Fig 1, illustrating that when transmission varies seasonally (Fig 1A) then any outbreak may be likely to fade out as soon as a season arrives that is not conducive to transmission (leading to a CER of zero; Fig 1B). However, even in that scenario, seasonal outbreaks may still lead to substantial numbers of cases (the TER may be larger than zero; Fig 1C).

**Figure 1.**
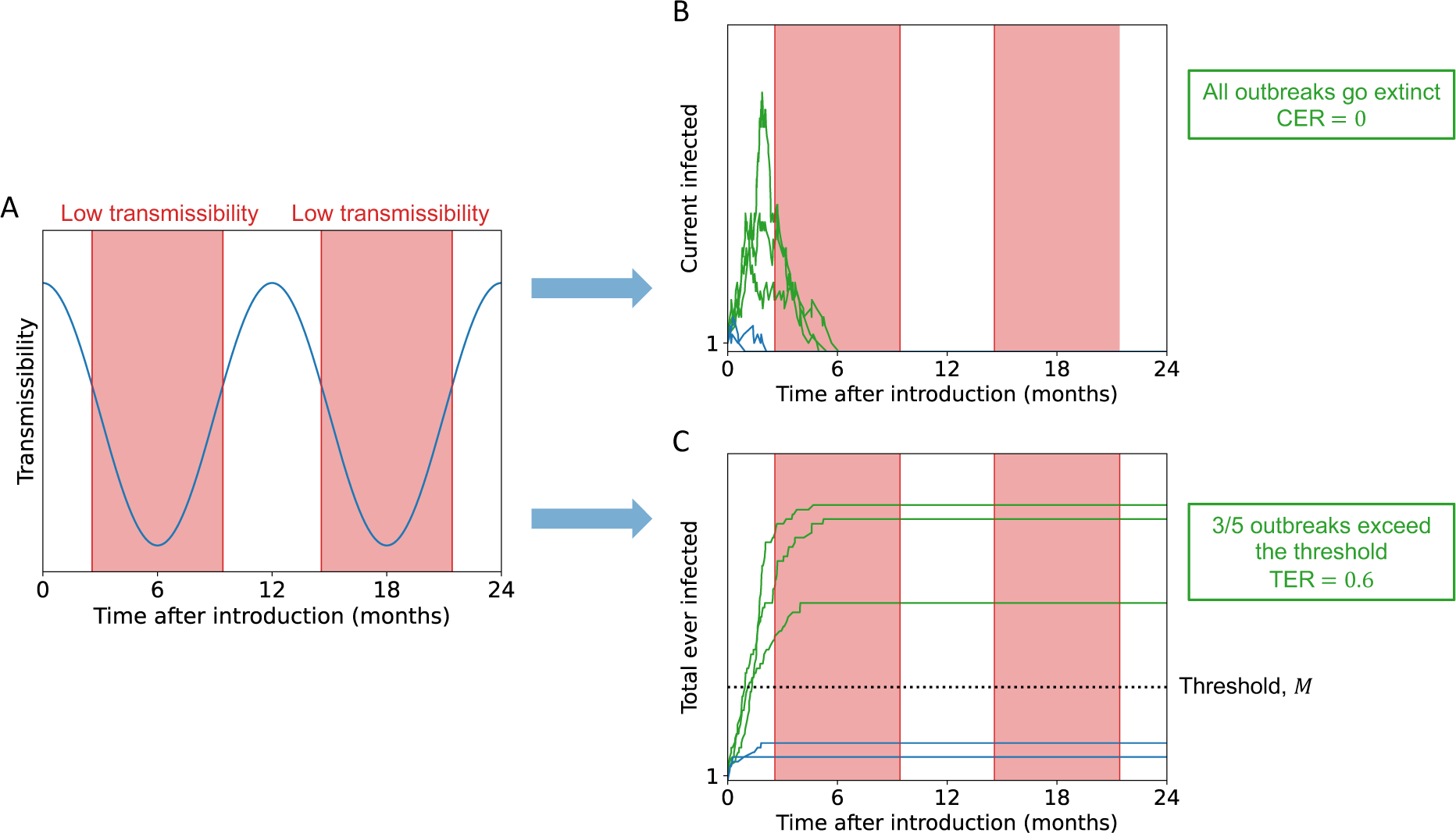
Schematic illustrating the difference in outbreak risk assessments for seasonal pathogens obtained using the CER and TER. A. Seasonal pathogen transmission comprises of periods of high and low transmissibility (low transmissibility periods, during which sustained pathogen transmission is impossible, are shaded in red). B. In the scenario considered here, all outbreaks go extinct during low transmissibility periods, leading to the CER taking the value zero. C. Despite all outbreaks going extinct, there is the potential for some outbreaks to generate a substantial number of cases. In this illustrative example, three out of every five outbreaks generate numbers of cases that exceed a pre-specified threshold, *M*, leading to a TER value of 0.6. In panels B and C, outbreaks that generate numbers of cases that exceed *M* are shown as green lines and those that do not are shown as blue lines.

First, we show how the TER can be calculated numerically (i.e., through the numerical solution of a system of equations, without requiring model simulations) for the stochastic SIR model with seasonally varying transmission. Then, we show how the TER can be calculated for more complex models using stochastic simulations by considering a stochastic host-vector model of Chikungunya virus transmission in northern Italy. When transmission is possible all year round, the TER and CER can give similar estimates. However, for both models, when there are substantial periods of the year during which sustained transmission is not possible, the difference between outbreak risk estimates arising from these two metrics can be large. For Chikungunya virus, which is spread by *Aedes albopictus*, there are long periods of the year in northern Italy during which vector abundance is too low for virus transmission [8]. Consequently, the CER is zero, yet major outbreaks due to local transmission can sometimes occur, depending on the precise definition of a “major outbreak” used. Since a policy-maker can choose a practically relevant threshold when estimating the TER, it is a useful quantity to consider when quantifying seasonal outbreak risks as an aid for public health policy making.

## 2. Methods

### 2.1 Epidemiological models

#### 2.1.1 SIR model

The ordinary differential equation (ODE) version of the Susceptible-Infectious-Removed (SIR) model with time-dependent infection and removal rates is:

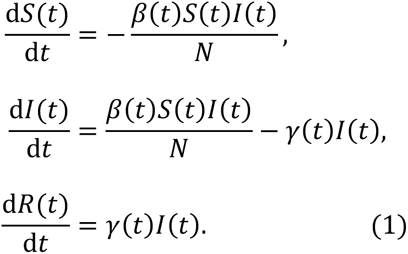

In this model, *S*(*t*) is the number of individuals who are susceptible to the pathogen at time *t*, *I*(*t*) is the number of infectious individuals, and *R*(*t*) is the number of removed individuals (including those who have recovered and become immune and those who have died). The total population size, *S*(*t*) + *I*(*t*) + *R*(*t*) = *N*, is constant under this model. In our analyses, the analogous stochastic model is considered, and simulations are run using a modified version of the Gillespie direct method [46] in which time-dependent rates are accounted for [29,47,48] (Text S1.2, Algorithm 1). For this model, the instantaneous basic reproduction number is given by 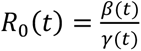.

Time *t* is measured in months and the infection rate is chosen to be periodic with a period of 12 months:

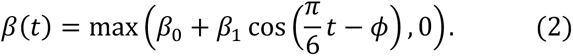

The removal rate is assumed to be constant (*γ*(*t*) = *γ*). We use these specific forms of the infection and removal rates in our analyses but our approach for computing the TER can be applied for any functions *β*(*t*) and *γ*(*t*) (the functions do not even need to be periodic). The parameter values used are shown in the captions to Figures 2-4.

**Figure 2.**
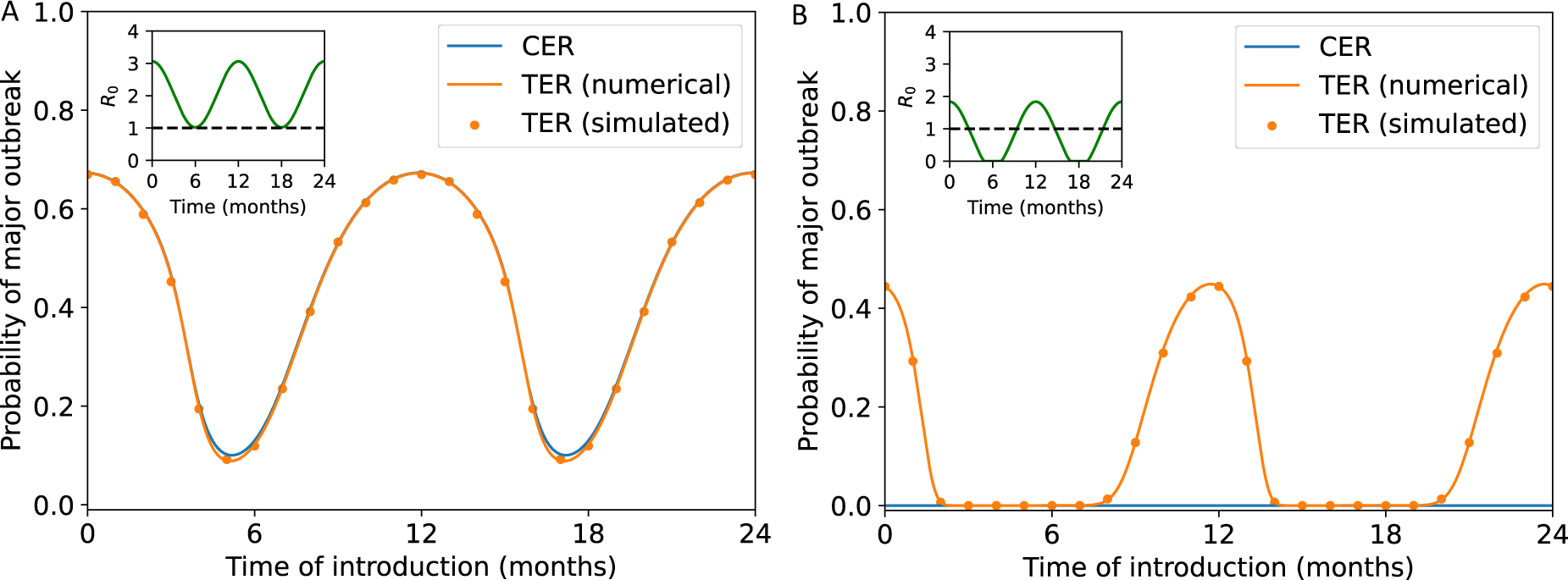
Comparison between calculated values of the CER and TER for the stochastic SIR model with seasonal transmission. A. The CER (obtained using equation (8) – blue line) and the TER (obtained by solving system of equations (11) numerically – orange line – and by running model simulations – orange dots) when sustained transmission is possible throughout the year (*β*_0_ = 10, *β*_1_ = 5 and *γ* = 4.9 month^-1^). B. Analogous results to panel A, but in a scenario in which sustained transmission can only occur for some of the year (*β*_0_ = 4, *β*_1_ = 5 and *γ* = 4.9 month^-1^). In both panels, a threshold of *M* = 100 was used when computing the TER and the overall population size was assumed to be *N* = 1,000 individuals. When we computed the TER using model simulations, we ran 10,000 simulations of the stochastic model (using the simulation approach described in Section 2.1.1) for each time of introduction considered. In both panels, the inset shows *R*_0_(*t*) = *β*(*t*)/*γ*(*t*) as a function of *t*.

#### 2.1.2 Chikungunya transmission model

We adapt the ODE model of Chikungunya virus transmission described by Guzzetta *et al.* [8,44]. Specifically, we separate the vector ecological dynamics from the host-vector epidemiological dynamics. The ecological model is given by:

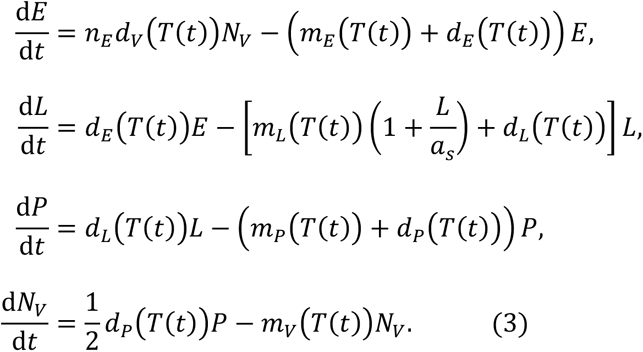

In this model, the population of vectors (*Ae. albopictus*) is split into eggs (*E*), larvae (*L*), pupae (*P*) and adults (*N*_*V*_). For notational convenience, we do not denote the dependence of these state variables on *t* in the equations above explicitly, although the number of vectors in each compartment of the model varies temporally. The factor of 1/2 in the equation for *N*_*V*_ reflects the fact that we only track adult female vectors, since male vectors do not spread the virus. The spatial scale of the model is assumed to be a single hectare (so that *N*_*V*_represents the number of adult female vectors in one hectare). The effect of overcrowded breeding sites on the larval mortality rate is determined by the overcrowding parameter, *a*_s_, which was fitted to vector capture data by Guzzetta *et al.* [8,44].

The temperature, *T*(*t*), is assumed to vary seasonally (i.e., with period 12 months):

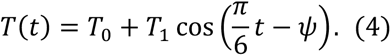

The values of *T*_0_, *T*_1_ and *ψ* were determined by fitting *T*(*t*) to daily mean temperature data (measured in Celsius) from Feltre, a town in northern Italy, separately for both 2014 and 2015 (data were obtained from MODIS satellite Land Surface Temperature measurements as detailed in [8]) using least squares estimation. In our analysis of the temperature data from 2014, time *t* = 0 corresponds to 1^st^ April 2014. In our analysis of the data from 2015, time *t* = 0 corresponds to 1^st^ April 2015.

We solve the ecological model (system of equations (3)) numerically to obtain *N*_*V*_(*t*). To facilitate straightforward computation of the CER (see below), we then fit a skewed and scaled Gaussian to the monthly values of *N*_*V*_(*t*) using least squares estimation, and use the resulting fitted version of *N*_*V*_(*t*) in all of our analyses. Again, we perform this fitting separately for 2014 and 2015. The fitted curve is of the form:

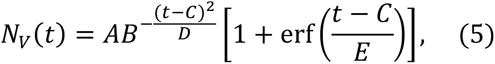

in which erf is the error function. By considering the deterministic version of the ecological model, we avoid running stochastic simulations of the ecological model, which would be computationally expensive due to the large number of events that would arise in that system.

Stochastic epidemiological dynamics are then simulated using a stochastic host-vector model. The analogous deterministic model to the stochastic model that we consider is:

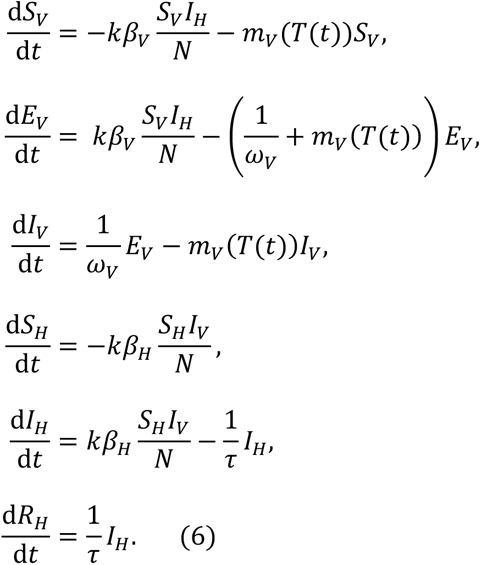

In this model, it is assumed that, after entering the *I*_*V*_ compartment, an adult female vector remains infectious for life. The temperature-dependent rates in systems of equations (3) and (6) are explicitly labelled as a function of temperature, *T*, which itself varies temporally. For definitions of each of the parameters in systems of equations (3) and (6), and the values used in our analyses (including functional forms of the temperature- dependent parameters), see Table S1.1. Unlike the total host population size, which remains constant (*S*_*H*_ + *I*_*H*_ + *R*_*H*_ = *N*), the vector population size, *N*_*V*_, varies with temperature and therefore varies temporally (equation (5)). The equation for the instantaneous basic reproduction number, *R*_0_(*t*), for this system is [8]:

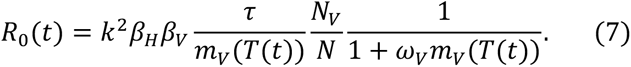

When we run stochastic simulations of system of equations (6), we again adapt the Gillespie direct method [46] (Text S1.2, Algorithm 2). We assume that transmission parameters take constant values within each day (given by their values at the start of the day). We are therefore able to use the Gillespie direct method within each day. At the end of each day, we compare the total vector population size, *S*_*V*_ + *E*_*V*_ + *I*_*V*_, with *N*_*V*_ (as determined by equation (5)). If *S*_*V*_ + *E*_*V*_ + *I*_*V*_ < *N*_*V*_, then we assume that new susceptible vectors are born (i.e., we increase *S*_*V*_) until *S*_*V*_ + *E*_*V*_ + *I*_*V*_ = *N*_*V*_. If instead *S*_*V*_ + *E*_*V*_ + *I*_*V*_ > *N*_*V*_, we select vectors uniformly at random to die until *S*_*V*_ + *E*_*V*_ + *I*_*V*_ = *N*_*V*_, since the per-vector death rates in system of equations (6) are equal for each of the *S*_*V*_, *E*_*V*_ and *I*_*V*_ compartments. By following this procedure, we simulate stochastic epidemiological dynamics while remaining consistent with the deterministic ecological dynamics (system of equations (3) and equation (5)).

### 2.2 Case Epidemic Risk (CER)

As described in the Introduction, a standard approach for estimating the probability of a major outbreak exists, involving the assumptions that infections occur according to a branching process and a constant supply of susceptible individuals is available for each infectious host to infect. This approach has been used previously in the context of pathogens for which transmission parameters vary temporally (e.g., [29,34,40]). Here, we refer to the probability of a major outbreak calculated in this way as the CER, following the use of this terminology in our earlier work [29]. In this section, we describe how the CER can be calculated for the stochastic SIR model and the stochastic host-vector model of Chikungunya virus transmission.

#### 2.2.1 SIR model

For the stochastic SIR model, if a single infectious individual enters the host population at time *t*_0_, then the CER is given by [29,34,40]:

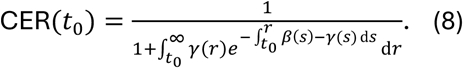

A derivation of this expression can be found in Section 2.3.1 of [29].

#### 2.2.2 Chikungunya transmission model

To compute the CER for the host-vector model of Chikungunya virus transmission, we use the method described in [29]. We denote the probability of a major outbreak occurring, if there are *i* infectious hosts, *j* exposed vectors and *k* infectious vectors at time *t*, by *p_ij_*_*k*_(*t*).

Assuming that the virus is introduced into the population at time *t*_0_by a single infectious host, then the CER is given by *p*_100_(*t*_0_). Calculation of the CER then involves solving the following system of ODEs:

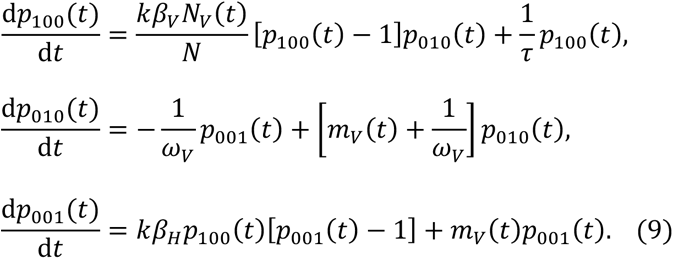

The first of these equations is derived in Text S1.3 with the derivation of the remaining two equations following an identical procedure. We numerically solve system of equations (9) using the Chebfun open source MATLAB software package [49], with periodic boundary conditions (*p*_100_(0) = *p*_100_(12), *p*_010_(0) = *p*_010_(12) and *p*_001_(0) = *p*_001_(12)). Chebfun requires the coefficients of *p*_100_, *p*_010_ and *p*_001_ on the right-hand- side of system of equations (9) to be provided in functional forms (as functions of *t*, rather than vectors of values), necessitating our decision to use a functional form for *N*_*V*_(*t*) (equation (5)).

### 2.3 Threshold Epidemic Risk (TER)

Here, we describe how the TER can be calculated for the stochastic SIR model and stochastic host-vector model of Chikungunya virus transmission. The TER represents the probability that, if a single infected individual (for the host-vector model, a single infected host) enters the population at time *t*_0_, an outbreak with more than a threshold number (denoted *M*) of infections follows. For the host-vector model, this threshold refers to the total number of host infections.

#### 2.3.1 SIR model

For the stochastic SIR model, we calculate the TER numerically, without resorting to model simulation. To do this, we choose a time, *t*_max_, that is longer than any outbreak could potentially be. We then denote the probability that the total number of infections exceeds *M* prior to time *t*_max_, given that there are *I*^∗^infectious individuals and *R*^∗^ removed individuals in the population at time *t*, by *q*_*M*_(*I*^∗^, *R*^∗^, *t*). In other words:

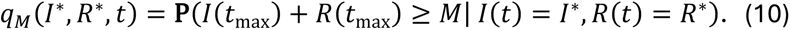

By choosing *t*_max_ to be longer than the timescale of any local outbreak, *q*_*M*_(*I*^∗^, *R*^∗^, *t*) is equivalent to the probability that at least *M* infections occur prior to outbreak extinction.

We discretise the time interval [0, *t*_max_] into *n* time steps, each of length Δ*t*, where Δ*t* is chosen to be small (by choosing *n* to be large) so that at most one event occurs in any time interval of length Δ*t*. By conditioning on the possible events occurring in the interval 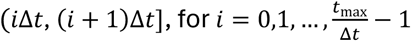, we obtain:

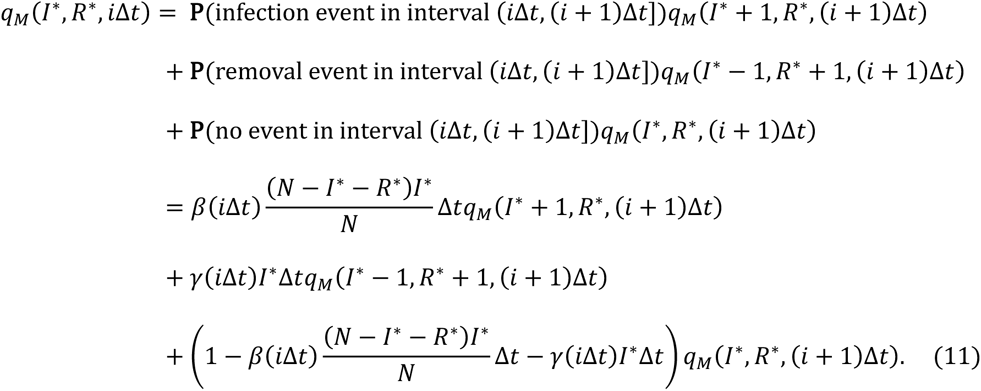

Since the outbreak will definitely have ended by time *t*_max_, we note that:

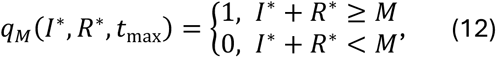

enabling us to solve system of equations (11) backwards in time to find the values of *q*_*M*_(*I*^∗^, *R*^∗^, *i*Δ*t*) for all values of *I*^∗^, *R*^∗^ and *i*. In other words, we first compute 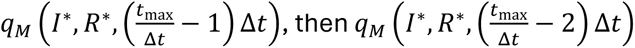, and so on. The TER, assuming that a single infectious individual is introduced to the host population at time *t*_0_, is then given by *q*_*M*_(1,0, *t*_0_).

We note that in principle it would be possible to rearrange system of equations (11) and take the limit Δ*t* → 0 to obtain a system of ODEs for *q*_*M*_(*I*^∗^, *R*^∗^, *t*). However, since we would then be required to discretise time to solve those ODEs numerically, we solve system of equations (11) directly as described above.

#### 2.3.2 Chikungunya transmission model

To compute the TER for the host-vector model, we use a simulation-based approach. Specifically, we repeatedly simulate the analogous stochastic model to system of equations (6), following the simulation procedure described in section 2.1.2. In each simulation, we start with a single infectious host in the population at time *t*_0_. The TER is then given by the proportion of model simulations in which *I*_*H*_ + *R*_*H*_ exceeds or equals *M* prior to pathogen extinction occurring.

## 3. Results

### 3.1 SIR model

To begin comparing the CER and TER, we calculated these quantities for the stochastic SIR model (the analogous stochastic model to system of equations (1)) with a seasonally varying infection rate (equation (2)). We first considered a scenario in which sustained transmission is possible all year round (*R*_0_(*t*) > 1 for all values of *t*), and set the threshold number of infections defining a “major outbreak” to be *M* = 100 when calculating the TER. We found that the TER matches the CER closely in that scenario (orange and blue lines in Fig 2A). Not only did we calculate the TER numerically using system of equations (11) (orange line in Fig 2A), but we also calculated the TER using repeated model simulation. To do this, we assumed that there was a single infected individual in the population at the time of pathogen introduction, *t*_0_ (i.e. *S*(*t*_0_) = *N* −1, *I*(*t*_0_) = 1 and *R*(*t*_0_) = 0), ran 10,000 simulations of the stochastic SIR model and then computed the proportion of simulations in which the number of infections exceeded or equalled *M* = 100 prior to outbreak extinction. We repeated this for a range of values of the time of introduction, *t*_0_ (orange dots in Fig 2A).

While the CER and TER matched closely when transmission was possible all year round (as was the case in previous studies in which the CER was calculated, e.g. [29]), we then went on to consider a second scenario in which sustained transmission is only possible for some of the year (Fig 2B). In that scenario, outbreaks with at least *M* = 100 infections were possible for some pathogen introduction times, leading to values of the TER that were greater than zero (orange line and dots in Fig 2B). However, since pathogen extinction always eventually occurred during time periods in which transmission was not possible, the CER took the value zero at all pathogen introduction times (blue line in Fig 2B).

Although we only considered a single introduced case in Fig 2, we also conducted a supplementary analysis in which we considered multiple pathogen introductions when calculating the TER (Fig S1.1).

We then explored the effect of the duration of time in the year for which sustained transmission is impossible on the mismatch between the CER and TER in more detail. Specifically, we considered varying the value of *β*_0_ and again calculated the CER and TER (Fig 3). As in Fig 2B, whenever transmission is not possible for long periods of the year, the CER always takes the value zero yet outbreaks with at least *M* = 100 infections may still occur (Fig 3A,B). If, however, there are periods of the year for which sustained transmission is impossible, but those periods are not very long, then CER values greater than zero but less than the TER can occur. This is because there is a chance that the pathogen survives in the host population across the periods during which conditions are not suitable for sustained transmission. We note that, even in those scenarios, if the duration of the year for which sustained transmission is impossible is not very short, then the CER is likely to suggest a lower outbreak risk than the TER, at least at some times of year (Fig 3C,D). Again, when sustained transmission is possible all year round, or is only impossible for very short periods, than the CER and TER match closely (Fig 3E,F).

**Figure 3.**
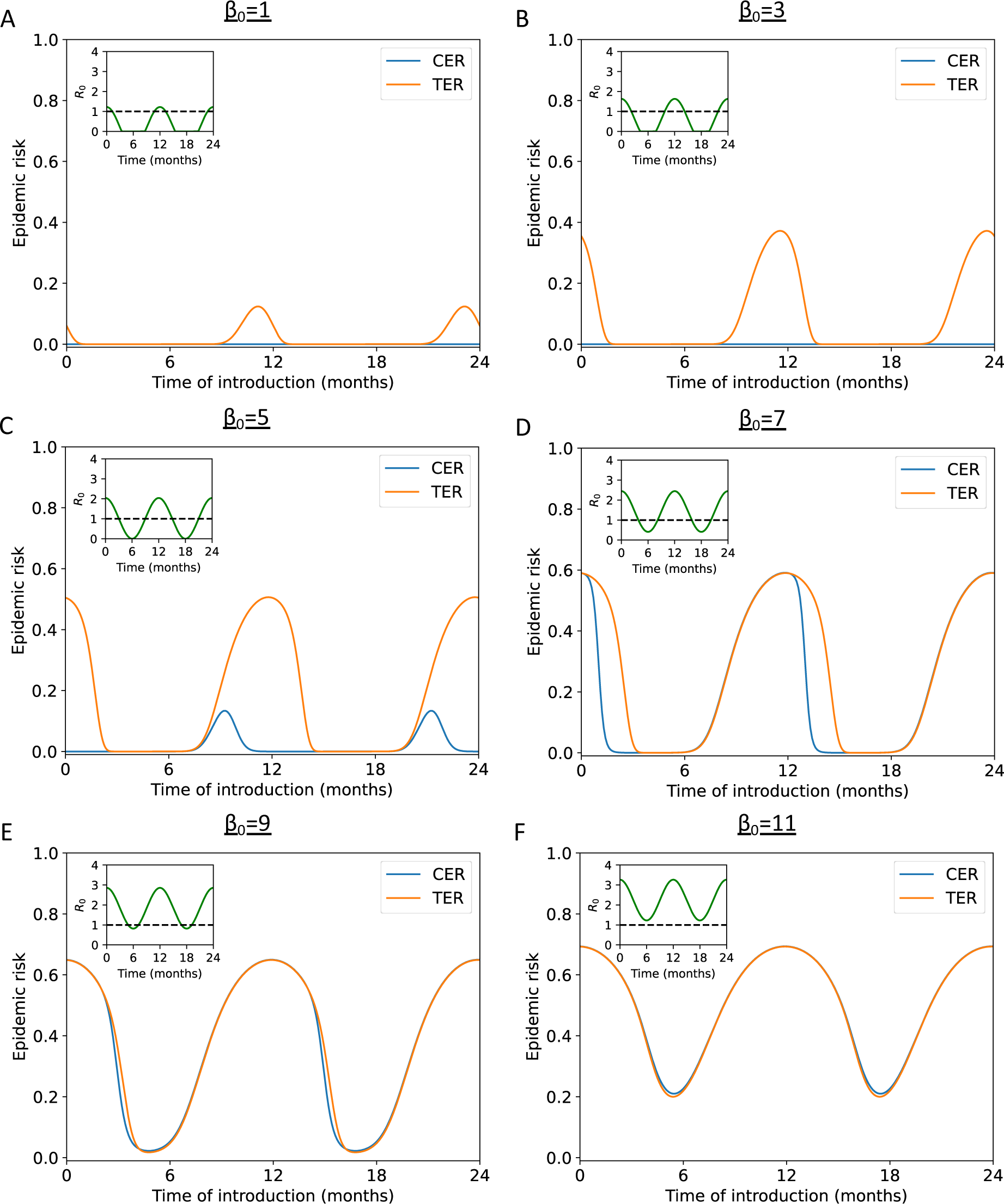
Comparison between calculated values of the CER and TER for the stochastic SIR model with seasonal transmission, for a range of values of *β*_0_. A. The CER (obtained using equation (8) – blue line) and the TER (obtained by solving systems of equations (11) numerically – orange line) when sustained transmission is only possible for a short period of the year (*β*_0_ = 1, *β*_1_ = 5 and *γ* = 4.9 month^-1^). B. Analogous results to panel A, but with *β*_0_ = 3. C. Analogous results to panel A, but with *β*_0_ = 5. D. Analogous results to panel A, but with *β*_0_ = 7. E. Analogous results to panel A, but with *β*_0_ = 9. F. Analogous results to panel A, but with *β*_0_ = 11. In all panels, a threshold of *M* = 100 was used when computing the TER and the overall population size was assumed to be *N* = 1,000 individuals. In all panels, the inset shows *R*_0_(*t*) = *β*(*t*)/*γ*(*t*) as a function of *t*.

We consider a similar analysis, but instead varying the extent of seasonality in the infection rate (*β*_1_), rather than *β*_0_, in Fig S1.2.

Having established that the TER provides a more appropriate characterisation of the risk posed by an invading seasonal pathogen than the CER, we considered the sensitivity of the TER to the precise threshold number of infections, *M*, chosen (Fig 4). Specifically, we considered both the value of the TER and the duration of the year for which the TER is above a particular value, *z* (in Fig 4, *z* = 0.1). For the transmission parameter values used in Fig 4 (*β*_0_ = 4, *β*_1_ = 5 and *γ* = 4.9 month^-1^), we found that the duration of the year in which the TER exceeds *z* = 0.1 was relatively similar for a range of dilerent values of *M*. For example, if *M* = 200 was used (corresponding to 20% of the host population), then the TER exceeded *z* = 0.1 for 5.36 months per year, whereas if instead *M* = 400 was used (corresponding to 40% of the host population), then the TER exceeded *z* = 0.1 for 4.60 months per year. We repeated this analysis for different values of *z* in Fig S1.3, and found similar results.

**Figure 4.**
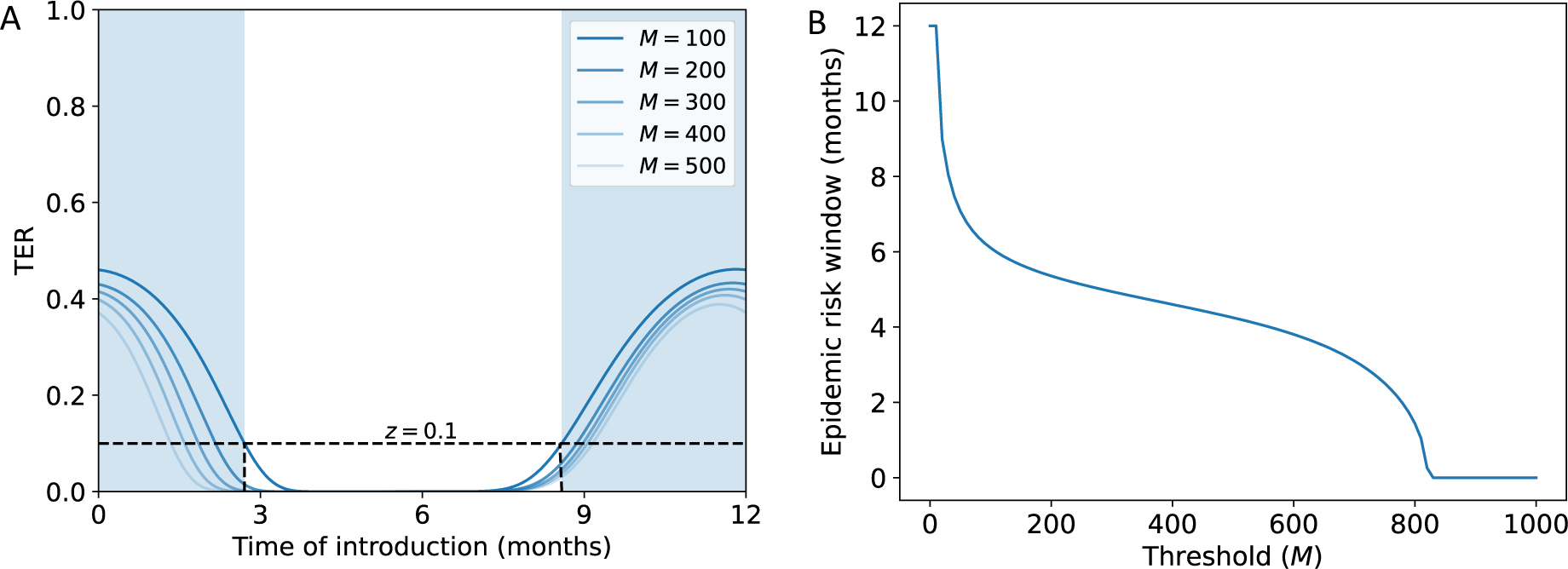
Sensitivity of the TER to the value of *M* chosen for the stochastic SIR model with seasonal transmission. A. The TER (obtained by solving systems of equations (11) numerically) for a range of different values of the threshold number of infections, *M*. The blue shaded region shows the period of the year for which the TER exceeds *z* = 0.1 for the baseline value of *M* = 100. B. The duration of the year for which the TER exceeds *z* = 0.1, shown as a function of *M*. In both panels, *β*_0_ = 4, *β*_1_ = 5 and *γ* = 4.9 month^-1^. The overall population size was assumed to be *N* = 1,000 individuals.

### 3.2 Chikungunya transmission model

To demonstrate the application of our framework for inferring the risk posed by an invading seasonal pathogen to a real-world case study, we estimated the TER for chikungunya in the town of Feltre, Italy, using daily mean temperature data from 2014 and 2015. The risk that an imported case will initiate a local outbreak varies during the year in that setting due to the seasonal dynamics of the *Ae. albopictus* vector population.

First, we fitted equation (4) to the temperature data from Feltre from 2014 (Fig S1.4A) and 2015 (Fig S1.4B). We then used these fitted temperature values to determine the number of adult female vectors per hectare throughout the year, initially by numerically solving system of equations (3) to obtain the number of adult female vectors at the start of each month (blue dots in Fig S1.4C,D) and then by fitting equation (5) to those monthly values (blue lines in Fig S1.4C,D). Finally, we computed the TER in 2014 (Fig 5A) and 2015 (Fig 5B) using model simulations, for a range of different values of the threshold number of infections defining a major outbreak, *M*. In addition to plotting the TER, we computed the CER and found that the CER was zero throughout each year due to the inevitable extinction of the pathogen during seasons in which environmental conditions are not conducive to transmission. Specifically, outside the summer months, low temperatures drive the vector population size down to a low level, making sustained transmission of Chikungunya impossible. This again highlights the importance of using the TER, rather than the CER, to quantify seasonal outbreak risks.

**Figure 5.**
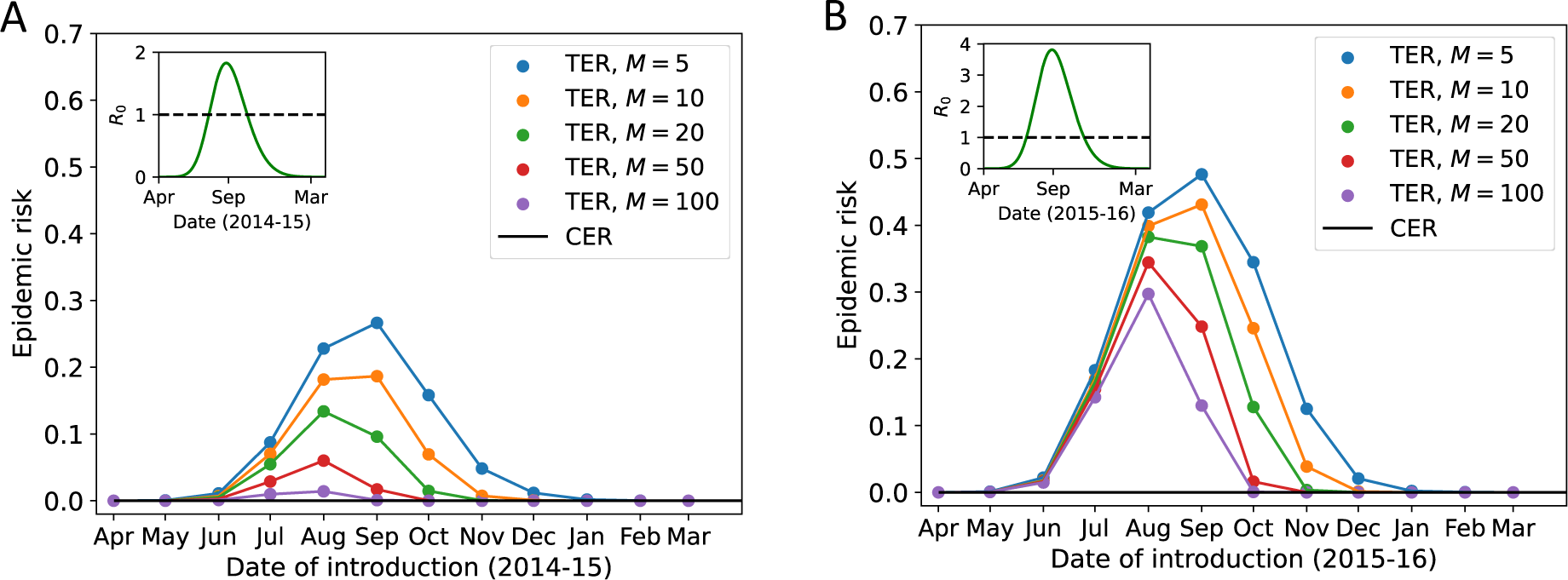
Calculation of the TER for chikungunya in Feltre, northern Italy, in 2014 and 2015. A. The TER for 2014 (and early 2015), shown for a range of values of the threshold number of infections, *M*. The CER is also shown (obtained using system of equations (9) – black line). B. Analogous to panel A, but for 2015 (and early 2016). In both panels, to compute the CER we ran 10,000 simulations of the stochastic model (using the simulation approach described in section 2.1.2) for each date of introduction considered, and the host population size was assumed to be *N* = 5,000 individuals (based on the population density in Feltre [44], this corresponds to an area of 80 Ha; the numbers of adult female vectors were also scaled up from their per Ha values shown in Fig S1.4C,D). In both panels, the inset shows *R*_0_(*t*) as a function of *t* (equation (7)).

## 4. Discussion

For many infectious diseases, quantifying the risk that imported cases will initiate a “major outbreak” driven by local transmission is of vital importance for public health policy. This is especially pertinent for seasonal pathogens that are not present at certain times of year, since pathogen reintroduction leading to sustained local transmission is necessary for large numbers of cases to arise. Identification of high risk locations and time periods allows policy-makers to target surveillance and control interventions appropriately.

Previous studies have provided a range of methods for calculating the probability of a major outbreak. For directly transmitted pathogens for which the parameters governing transmission do not vary temporally, the probability of a major outbreak starting from a single infected individual can be approximated by 1 − 1/*R*_0_ (whenever *R*_0_ > 1), in which *R*_0_ is the basic reproduction number of the pathogen [50]. When transmission parameter values vary temporally, a more complex calculation is required, and an established method [29,34–41] gives rise to the quantity that we term the CER here. The CER has previously been calculated for a range of models, including the stochastic SIR model with seasonally varying transmission [29,34–41], host-vector models [29,40], and models that account for varying susceptible population sizes [15,20].

As we have shown, when sustained transmission is possible all year around, the CER provides a useful measure of the risk that an introduced case will initiate local transmission (Figs 2A, 3F). However, when sustained transmission cannot occur for substantial periods of the year (e.g. over winter, as is the case for vector-borne pathogens in temperate climates), then the CER can underestimate the true risk of a substantial outbreak occurring, including scenarios in which outbreaks with large numbers of cases can occur yet the CER takes the value zero (Figs 2B, 3A,B). For this reason, we have proposed a new quantity that can be calculated for assessing the probability of a major outbreak. Specifically, the TER represents the probability that introduced cases initiate an outbreak with more than *M* infections prior to outbreak extinction.

The risk of an outbreak with more than a threshold number of cases occurring has been considered before for pathogens for which transmission parameters remain constant in time [30]. In that scenario, the TER tends to match classic estimates for the probability of a major outbreak for a range of values of *M*, at least when *R*_0_ is sufficiently larger than one. In contrast, for seasonal pathogens, as noted above there are scenarios in which substantial outbreaks can occur but the CER is zero, demonstrating a clear mismatch between standard estimates for the probability of a major outbreak and estimates with clear practical meaning such as the TER.

Although we found that the precise value of *M* chosen did not always alect the calculated value of the TER substantially (Fig 4), the value of *M* may be chosen by policy-makers in a context specific fashion. For example, for a pathogen such as the dengue virus in Italy, even relatively small outbreaks would be considered substantial. Since 2010, each dengue outbreak in Europe has resulted in fewer than 100 reported cases [51]. Therefore, even outbreaks with tens of cases might be considered large in that setting, suggesting that a value of *M* of that order of magnitude might be appropriate. In practice, if mathematical modellers undertake calculation of the TER, then we contend that this should be done for any specific outbreak in consultation with policy-makers, to ensure that an appropriate value of *M* is used. Alternatively, the TER could be computed for a range of values of *M*, so that estimates of the risk of outbreaks of a range of dilerent sizes are obtained.

As we showed by applying our approach to the case-study of chikungunya in northern Italy (Fig 5), the methodology presented here is particularly relevant in the context of vector-borne diseases in locations that experience seasonal outbreaks. Going forwards, the risk of vector-borne disease outbreaks is expected to increase in some locations due to climate change [52,53]. Calculation of the TER across a range of places and at different times of year can provide insights into changes in the spatio-temporal risk of outbreaks and support the adoption of preventive measures [44].

In addition to demonstrating that the CER does not provide an appropriate assessment of the risk of seasonal outbreaks in a real-world scenario, three features are particularly noticeable from our TER calculations in Fig 5. First, relatively small differences in temperature between years (Fig S1.4A,B) can drive more substantial differences in the vector population size (Fig S1.4C,D), and therefore in the risk posed by outbreaks (Fig 5). Second, the choice of value of *M* affects the time of pathogen introduction at which the TER is maximised. Specifically, larger values of *M* require longer outbreaks for the threshold number of infections to be exceeded. As a result, larger values of *M* tend to lead to earlier peak values of the TER, in order for there to be sufficient time left in the transmission season for such large outbreaks to occur. Third, and relatedly, the level of variation in the TER between different values of *M* can change during the year. In Fig 5B, for example, early in the transmission season the TER was similar across the range of values of *M* considered. This is because simulated outbreaks tended to either fade out with few infections or large numbers of infections (more than 100) occurred. In contrast, later in the transmission season, because of the limited time period remaining until sustained transmission became impossible, outbreaks of a range of different sizes could arise. This highlights the need to consider the value of *M* used when calculating the TER carefully in some scenarios.

In summary, we have developed a novel framework for seasonal pathogens that can be used to compute the probability that an initial infected case (or cases) initiates a “major outbreak”. Rather than basing our approach on the mathematical theory of branching processes, which can lead to unrealistic assessments of seasonal outbreak risks, we calculate the TER (i.e., the probability that the number of infections will exceed a pre-specified threshold value) directly. For simple stochastic epidemic models that account for seasonality, the TER can be calculated numerically. For more complex models, the TER can be estimated using model simulations, enabling it to be determined for any epidemiological system for which repeated model simulation is possible. Going forwards, we hope that our flexible approach will be used by epidemiological modellers to obtain policy-relevant outbreak risk assessments for a range of seasonal pathogens.

## Data Availability

All data are available online

https://github.com/KayeARK/Quantifying_Epidemic_Risks

## AUTHORS’ CONTRIBUTIONS

ARK – conceptualization, methodology, formal analysis, investigation, software, validation, writing – original draft, writing – review and editing.

GG – methodology, writing – review and editing. MJT – supervision, writing – review and editing.

RNT – conceptualization, methodology, project administration, supervision, writing – original draft, writing – review and editing.

## ACKNOWLEDGEMENTS

Thanks to members of the Zeeman Institute for Systems Biology and Infectious Disease Epidemiology Research at the University of Warwick, the Wolfson Centre for Mathematical Biology at the University of Oxford and the Centre for Health Emergencies at the Bruno Kessler Foundation for useful discussions about this research.

## DATA SHARING

All data generated or analysed during this study, including computing code for reproducing our results, are available at: www.github.com/KayeARK/Quantifying_Epidemic_Risks

