## Supplementary material for "Quantifying infectious disease epidemic risks: A practical approach for seasonal pathogens"

### Supplementary Text

#### Text S1.1 – Parameters of the Chikungunya transmission model

The parameters of the model of Chikungunya virus transmission are listed in Table S1.1, along with the assumed parameter values. For the temperature-dependent parameter values, a functional form is provided (based on the temperature, $T$).

**Table S1.1. Parameters of the *Ae. albopictus* ecological model and the Chikungunya transmission model (systems of equations (3) and (6) in the main text).** Graphical representations of parameters that are temperature-dependent are shown in Poletti *et al.* [1]. Relevant references from which the assumed values and functional forms were obtained are shown in the final column.

| Parameter | Interpretation | Value | Units | Reference |
|---|---|---|---|---|
| $n_E$ | Average number of eggs per adult female oviposition | 60 | - | [1–3] |
| $d_E(T)$ | Development rate from egg to larva | $\frac{1}{6.9 - 4e^{-\left(\frac{T-20}{4.1}\right)^2}}$ | $\text{days}^{-1}$ | [1–3] |
| $d_L(T)$ | Development rate from larva to pupa | $\frac{1}{0.12T^2 - 6.6T + 98}$ | $\text{days}^{-1}$ | [1–3] |
| $d_P(T)$ | Development rate from pupa to adult | $\frac{1}{0.027T^2 - 1.7T + 27.7}$ | $\text{days}^{-1}$ | [1–3] |
| $d_V(T)$ | Rate of egg deposition for female adults | $\frac{1}{0.046T^2 - 2.77T + 45.3}$ | $\text{days}^{-1}$ | [1–3] |
| $m_E(T)$ | Egg mortality rate | $506 - 506e^{-\left(\frac{T-25}{27.3}\right)^6}$ | $\text{days}^{-1}$ | [1–3] |
| $m_L(T)$ | Larval mortality rate | $0.029 + 858e^{T-43.4}$ | $\text{days}^{-1}$ | [1–3] |
| $m_P(T)$ | Pupal mortality rate | $0.021 + 37e^{T-36.8}$ | $\text{days}^{-1}$ | [1–3] |
| $m_V(T)$ | Adult vector mortality rate | $0.031 + 95820e^{T-50.4}$ | $\text{days}^{-1}$ | [1–3] |
| $k$ | Adult vector biting rate | 0.09 | $\text{days}^{-1}$ | [1,2] |
| $\beta_V$ | Probability of vector infection from a single blood meal from an infectious host | 0.77 | - | [2] |

| | | | | |
|---|---|---|---|---|
| $\beta_H$ | Probability of host infection from a single blood meal from an infectious vector | 0.70 | - | [2] |
| $\omega_V$ | Extrinsic incubation period | 2.5 | days | [2] |
| $\tau$ | Time until host recovery | 4.5 | days | [2] |
| $a_s$ | Overcrowding term | 44.5 (2014), 32.6 (2015) | - | [2] |

### Text S1.2 - Supplementary details about the algorithms used for outbreak simulations

#### Algorithm 1 – Seasonal stochastic SIR model

Two possible events can occur in the stochastic SIR model. These events, and the rates at which they occur, are shown in Table S1.2.

**Table S1.2. Possible events in the stochastic SIR model.**

| **Event** | **Rate** |
|---|---|
| A randomly chosen susceptible individual becomes infected | $\frac{\beta(t)S(t)I(t)}{N}$ |
| A randomly chosen infected individual is removed | $\gamma(t)I(t)$ |

A single realisation of the stochastic SIR model can be simulated using the following algorithm:

1. Set the initial time $t$ and the values of $S(t), I(t)$ and $R(t)$.
2. Steps 2-3 should be repeated while the outbreak is still ongoing (i.e. $I(t) > 0$) or until the user chooses to stop the simulation. Calculate the time of the next event, $t+\tau$, using the expression

   $$\int_t^{t+\tau} \frac{\beta(s)S(s)I(s)}{N} + \gamma(s)I(s)\mathrm{d}s = -\ln(r_1),$$

   where $r_1$ is a random number sampled from a uniform distribution on $(0,1)$.

3. Determine whether the next event is an infection event or a removal event. To do this, sample a second random number ($r_2$) from a uniform distribution on $(0,1)$. If

$$r_2 < \frac{\beta(t+\tau)S(t)I(t)/N}{\beta(t+\tau)S(t)I(t)/N + \gamma(t+\tau)I(t)},$$

then the next event is an infection event; set $S(t+\tau) = S(t) - 1$, $I(t+\tau) = I(t) + 1$, and $R(t+\tau) = R(t)$. If instead the inequality above is not satisfied, then the next event is a removal event; set $S(t+\tau) = S(t)$, $I(t+\tau) = I(t) - 1$, and $R(t+\tau) = R(t) + 1$. Update the current time, $t$.

Algorithm 2 – Seasonal stochastic Chikungunya transmission model

As described in the main text, to simulate the Chikungunya transmission model, we begin by solving the deterministic ecological model (system of equations (3) in the main text) numerically. Following the approach of Guzzetta *et al.* [2,4], the ecological model is initialised on 1st April of the respective years with 10,000 eggs and no individuals in any other compartment. We then fit equation (5) in the main text to the output from system of equations (3) to obtain a time series describing the number of adult female vectors each day, $N_V(t)$.

We then proceed by simulating the epidemiological component of the system (the analogous stochastic model to system of equations (6) in the main text). Within each day, transmission parameter values are assumed to remain constant. Seven different types of event can occur in the simulations within each day (Table S1.3).

**Table S1.3. Possible events within each day in the stochastic Chikungunya transmission model.**

| Event | Rate |
|---|---|
| A randomly chosen susceptible vector dies | $m_V(T(t))S_V$ |
| A randomly chosen susceptible vector becomes exposed | $k\beta_V \frac{S_V I_H}{N}$ |

| | |
|---|---|
| A randomly chosen exposed vector dies | $m_V(T(t))E_V$ |
| A randomly chosen exposed vector becomes infectious | $\frac{1}{\omega_V}E_V$ |
| A randomly chosen infectious vector dies | $m_V(T(t))I_V$ |
| A randomly chosen susceptible host becomes infectious | $k\beta_H\frac{S_H I_V}{N}$ |
| A randomly chosen infectious host is removed | $\frac{1}{\tau}I_H$ |

The stochastic host-vector model is then simulated using the following steps:

1. Set the initial time $t$, and the values of $S_V(t)$, $E_V(t)$, $I_V(t)$, $S_H(t)$, $I_H(t)$ and $R_H(t)$. $S_V(t)$ is set to be $N_V(t)$ (from the deterministic ecological model), $S_H(t)$ is set to be the number of people living within the simulation area, and $I_H(t)$ is set to be one (this is the initial infection). All other compartments are initialised with zero individuals.
2. Steps 2-3 should be repeated while the outbreak is still ongoing (i.e. $E_V + I_V + I_H > 0$) or until the user chooses to stop the simulation. Propose the next event time, $t + \tau$, using the expression

$$\tau = -\frac{\ln r_1}{\theta(t)},$$

where $\theta(t) = m_V\big(T(t)\big)S_V(t) + k\beta_V\frac{S_V(t)I_H(t)}{N} + m_V(T(t))E_V(t) + \frac{1}{\omega_V}E_V(t) + m_V(T(t))I_V(t) + k\beta_H\frac{S_H(t)I_V(t)}{N} + \frac{1}{\tau}I_H(t)$ and $r_1$ is a random number sampled from a uniform distribution on $(0,1)$.

3. Then, do one of the following, depending on the proposed next event time $(t + \tau)$:
   - If this proposed time $t + \tau$ is not in the same day as time $t$ (i.e. if $\lfloor t + \tau \rfloor - \lfloor t \rfloor > 0$), then do not perform any event and instead update the time to be the end of the original day (i.e. update $t$ to $\lceil t \rceil$). Update the vector population size according to the deterministic ecological model (i.e. $N_V(t)$). Let $\sigma(t) = N_V(t) - S_V(t) - E_V(t) -$

$I_V(t)$. If $\sigma(t) > 0$, then add $\sigma(t)$ individuals to $S_V(t)$. If $\sigma(t) < 0$, then remove $-\sigma(t)$ individuals from $S_V(t)$, $E_V(t)$ or $I_V(t)$ (with each vector to remove chosen uniformly at random from those compartments).

- If instead the proposed time $t + \tau$ is in the same day as $t$ (i.e. if $\lfloor t + \tau \rfloor - \lfloor t \rfloor = 0$), do not update the vector population size. Determine the type of the event occurring at time $t + \tau$ (since transmission parameters are assumed to be constant within each day, this is equivalent to deploying the Gillespie direct method within each day). To do this, define $\theta_i(t)$ to be the sum of the first $i$ terms of $\theta(t)$, so that $\theta_1(t) = m_V\big(T(t)\big)S_V(t)$, $\theta_2(t) = m_V(T(t))S_V(t) + k\beta_V \frac{S_V(t)I_H(t)}{N}$, and so on. Then, sample a second number ($r_2$) from a uniform distribution on $(0,1)$, and perform one of the following events.
    - If $r_2 < \frac{\theta_1(t)}{\theta(t)}$, then the next event is the death of a susceptible vector; set $S_V(t+\tau) = S_V(t) - 1$, $E_V(t+\tau) = E_V(t)$, $I_V(t+\tau) = I_V(t)$, $S_H(t+\tau) = S_H(t)$, $I_H(t+\tau) = I_H(t)$, and $R_H(t+\tau) = R_H(t)$.
    - If instead, $\frac{\theta_1(t)}{\theta(t)} < r_2 < \frac{\theta_2(t)}{\theta(t)}$, then the next event is a susceptible vector becoming exposed; $S_V(t+\tau) = S_V(t) - 1$, $E_V(t+\tau) = E_V(t) + 1$, $I_V(t+\tau) = I_V(t)$, $S_H(t+\tau) = S_H(t)$, $I_H(t+\tau) = I_H(t)$, and $R_H(t+\tau) = R_H(t)$.
    - If instead, $\frac{\theta_2(t)}{\theta(t)} < r_2 < \frac{\theta_3(t)}{\theta(t)}$, then the next event is the death of an exposed vector; $S_V(t+\tau) = S_V(t)$, $E_V(t+\tau) = E_V(t) - 1$, $I_V(t+\tau) = I_V(t)$, $S_H(t+\tau) = S_H(t)$, $I_H(t+\tau) = I_H(t)$, and $R_H(t+\tau) = R_H(t)$.
    - If instead, $\frac{\theta_3(t)}{\theta(t)} < r_2 < \frac{\theta_4(t)}{\theta(t)}$, then the next event is the transition of an exposed vector into an infectious vector; set $S_V(t+\tau) = S_V(t)$, $E_V(t+\tau) = E_V(t) - 1$,

$I_V(t+\tau) = I_V(t) + 1$, $S_H(t+\tau) = S_H(t)$, $I_H(t+\tau) = I_H(t)$, and $R_H(t+\tau) = R_H(t)$.

- If instead, $\frac{\theta_4(t)}{\theta(t)} < r_2 < \frac{\theta_5(t)}{\theta(t)}$, then the next event is the death of an infectious vector; set $S_V(t+\tau) = S_V(t)$, $E_V(t+\tau) = E_V(t)$, $I_V(t+\tau) = I_V(t) - 1$, $S_H(t+\tau) = S_H(t)$, $I_H(t+\tau) = I_H(t)$, and $R_H(t+\tau) = R_H(t)$.
- If instead, $\frac{\theta_5(t)}{\theta(t)} < r_2 < \frac{\theta_6(t)}{\theta(t)}$, then the next event is the infection of a susceptible host; set $S_V(t+\tau) = S_V(t)$, $E_V(t+\tau) = E_V(t)$, $I_V(t+\tau) = I_V(t)$, $S_H(t+\tau) = S_H(t) - 1$, $I_H(t+\tau) = I_H(t) + 1$, and $R_H(t+\tau) = R_H(t)$.
- If instead, $\frac{\theta_6(t)}{\theta(t)} < r_2 < 1$, then the next event is the removal of an infectious host; set $S_V(t+\tau) = S_V(t)$, $E_V(t+\tau) = E_V(t)$, $I_V(t+\tau) = I_V(t)$, $S_H(t+\tau) = S_H(t)$, $I_H(t+\tau) = I_H(t) - 1$, and $R_H(t+\tau) = R_H(t) + 1$.

Then update the current time, $t$ (i.e. set $t$ to be $t + \tau$).

**Text S1.3 – Derivation of the CER for the host-vector model**

We derive the CER for the host-vector model, considering a scenario in which a single infectious host enters the population at time $t = t_0$. To do this, we denote the probability of a major outbreak failing to occur starting from $i$ infectious hosts, $j$ exposed vectors and $k$ infectious vectors in the population at time $t = t_0$ by $q_{ijk}(t_0)$.

We begin by assuming that there is one infectious host and no exposed or infectious vectors in the population in order to write down an equation for the temporal evolution of $q_{100}$. We then consider the possible events in the next $\Delta t$ months (i.e. the time interval $[t_0, t_0 + \Delta t]$), where $\Delta t$ represents a very short time period so that at most a single event is possible. In that time period, the probability that a vector is infected (i.e. transitions from the susceptible compartment to the exposed compartment) is approximately $k\beta_V \frac{S_V I_H}{N} \Delta t$; the probability that the infectious host recovers is approximately $\frac{1}{\tau}\Delta t$; and the probability that no event occurs is approximately $1 - k\beta_V \frac{S_V I_H}{N} \Delta t - \frac{1}{\tau}\Delta t$. Applying the law of total probability gives

$$\begin{aligned}
q_{100}(t_0) \; &= \text{Prob}(\text{vector infection event occurs in } [t_0, t_0 + \Delta t]) \\
&\quad \times \text{Prob}(\text{no major outbreak} \mid \text{vector infection event occurs in } [t_0, t_0 + \Delta t]) \\
&\quad + \text{Prob}(\text{host recovery event occurs in } [t_0, t_0 + \Delta t]) \\
&\quad \times \text{Prob}(\text{no major outbreak} \mid \text{host recovery event occurs in } [t_0, t_0 + \Delta t]) \\
&\quad + \text{Prob}(\text{no event occurs in } [t_0, t_0 + \Delta t]) \\
&\quad \times \text{Prob}(\text{no major outbreak} \mid \text{no event occurs in } [t_0, t_0 + \Delta t])
\end{aligned}$$

$$\begin{aligned}
&= k\beta_V \frac{S_V I_H}{N} \Delta t q_{110}(t_0 + \Delta t) + \frac{1}{\tau}\Delta t q_{000}(t_0 + \Delta t) \\
&\quad + \left(1 - k\beta_V \frac{S_V I_H}{N} \Delta t - \frac{1}{\tau}\Delta t\right) q_{100}(t_0 + \Delta t).
\end{aligned}$$

Making the assumption that infection lineages are independent (so that $q_{110}(t_0 + \Delta t) = q_{100}(t_0 + \Delta t) q_{010}(t_0 + \Delta t)$), and noting that $q_{000}(t_0 + \Delta t) = 1$, $S_V = N_V$ and $I_H = 1$ at the beginning of the outbreak, then rearranging this expression and taking the limit $\Delta t \to 0$ gives

$$\frac{\mathrm{d}q_{100}(t_0)}{\mathrm{d}t_0} = -k\beta_V \frac{N_V}{N} q_{100}(t_0) q_{010}(t_0) - \frac{1}{\tau} + \left(k\beta_V \frac{N_V}{N} + \frac{1}{\tau}\right) q_{100}(t_0).$$

Denoting the probability of a major outbreak starting from $i$ infectious hosts, $j$ exposed vectors and $k$ infectious vectors in the population at time $t = t_0$ by $p_{ijk}(t_0) = 1 - q_{ijk}(t_0)$ and substituting this into the above equation gives the first equation in system of equations

(9) in the main text. An analogous approach (this time for $q_{010}$ and $q_{001}$; i.e. starting from either a single exposed vector or from a single infectious vector) is used to derive the remaining two equations in system of equations (9) in the main text.

### **Supplementary Figures**

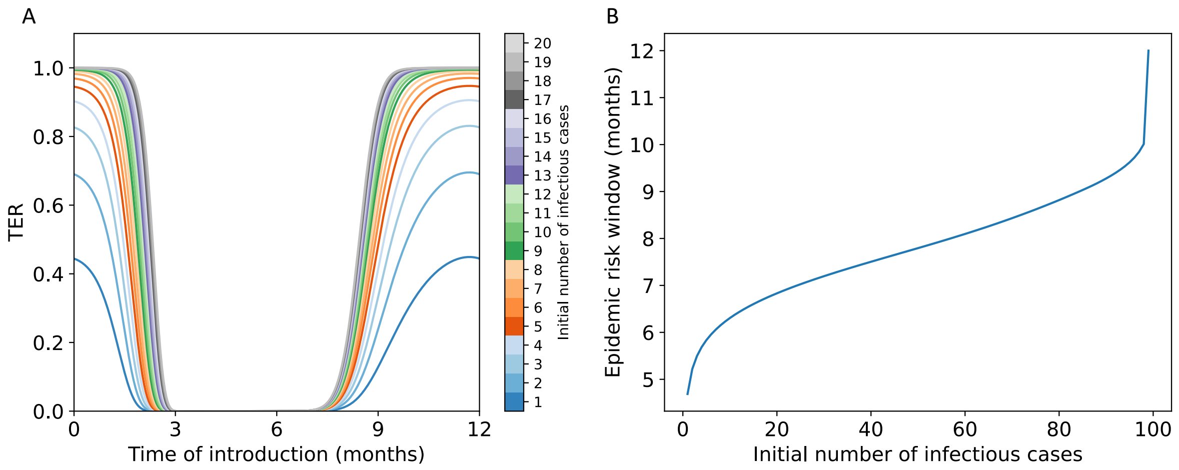


**Figure S1.1. Dependence of the TER on the initial number of infected individuals, for the stochastic SIR model with seasonal transmission.** A. The TER for different initial numbers of infectious individuals (obtained by solving systems of equations (11) in the main text numerically). B. The duration of the year for which the TER exceeds $z = 0.1$, for different initial numbers of infectious individuals. In both panels, a threshold of $M = 100$ was used when computing the TER and the overall population size was assumed to be $N = 1{,}000$ individuals. Parameter values used: $\beta_0 = 4$, $\beta_1 = 5$ and $\gamma = 4.9$ month$^{-1}$.

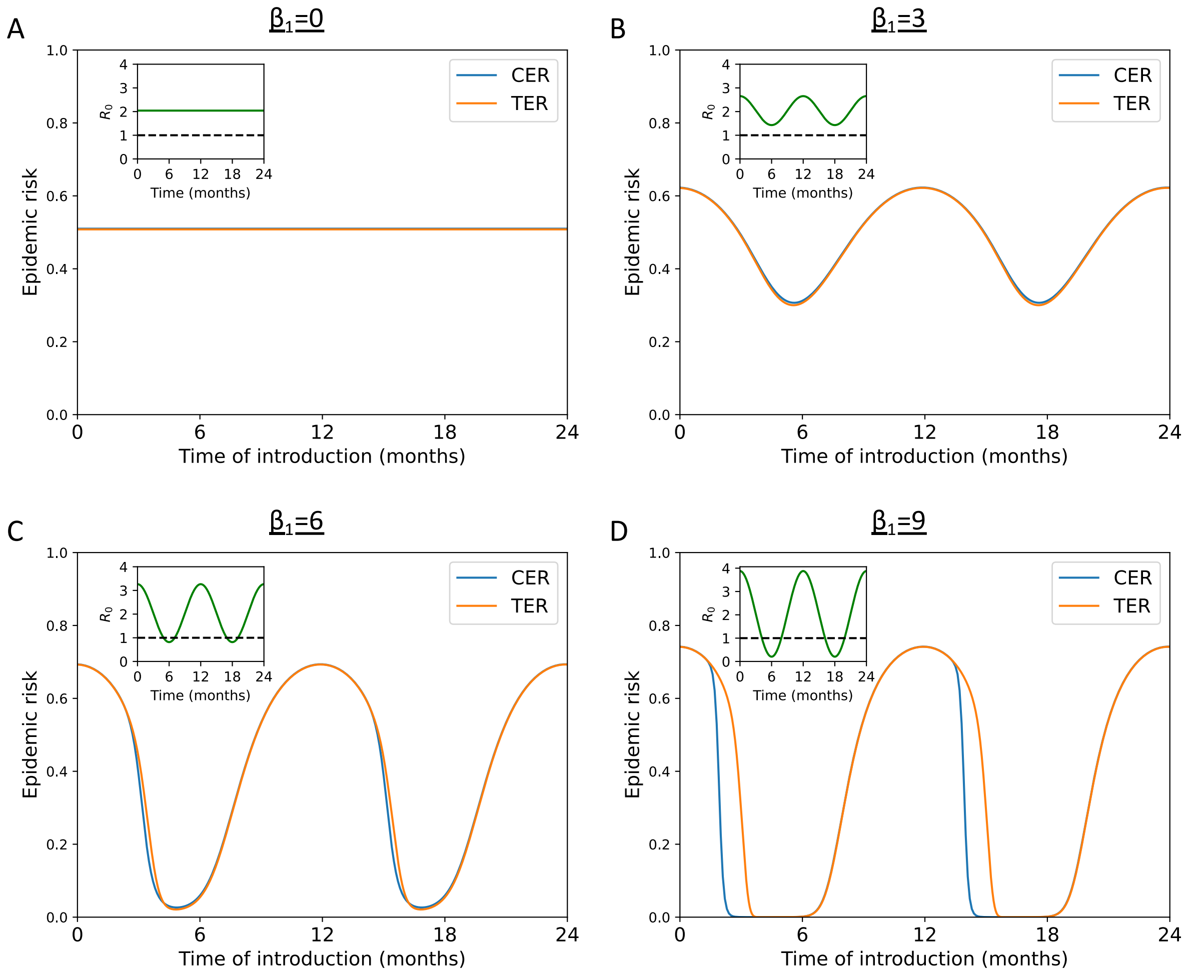


**Figure S1.2. Comparison between calculated values of the CER and TER for the stochastic SIR model with seasonal transmission, for a range of values of $\boldsymbol{\beta_1}$.** A. The CER (equation (8) in the main text – blue line) and the TER (obtained by solving systems of equations (11) in the main text numerically – orange line) when $\beta_0 = 10$, $\beta_1 = 0$ and $\gamma = 4.9$ month$^{-1}$. B. Analogous results to panel A, but with $\beta_1 = 3$. C. Analogous results to panel A, but with $\beta_1 = 6$. D. Analogous results to panel A, but with $\beta_1 = 9$. In all panels, a threshold of $M = 100$ was used when computing the TER and the overall population size was assumed to be $N = 1{,}000$ individuals. In all panels, the inset shows $R_0(t) = \beta(t)/\gamma(t)$ as a function of $t$.

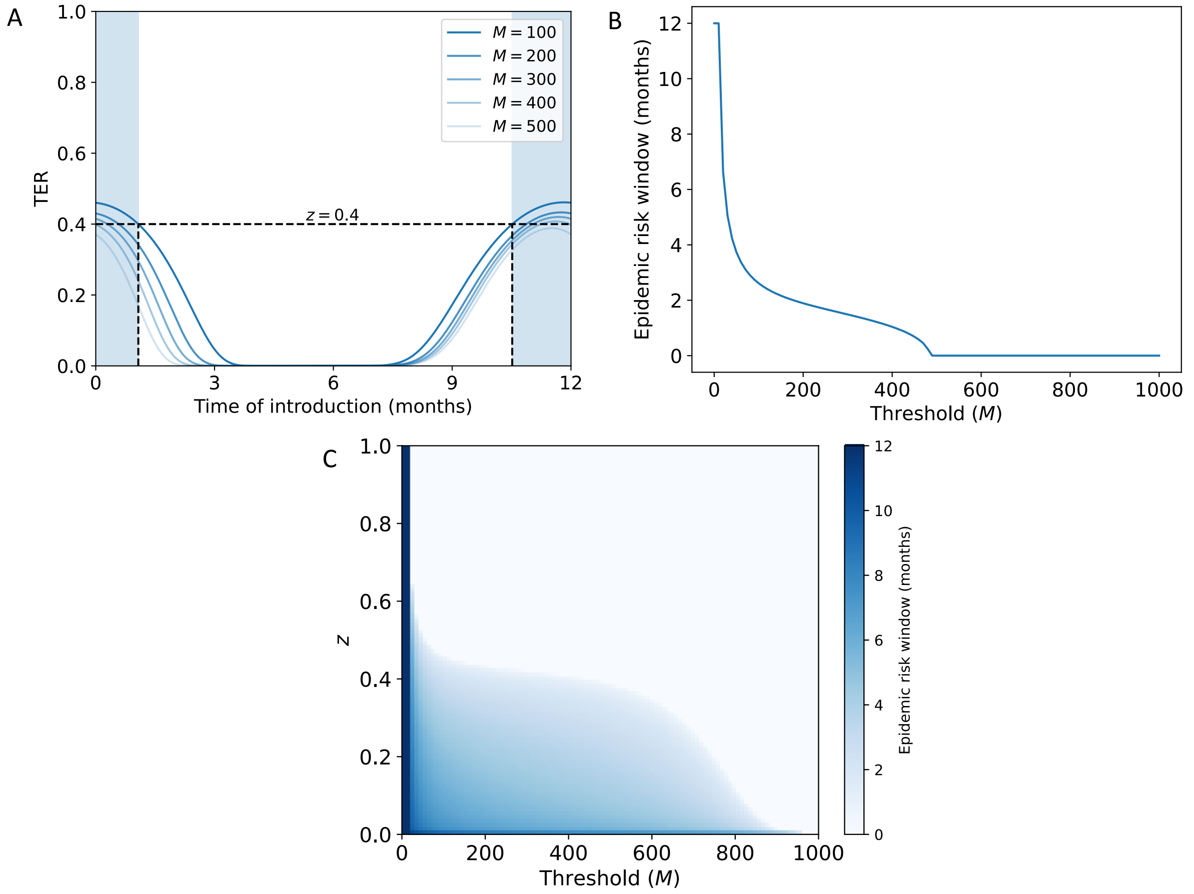


**Figure S1.3. Proportion of the year for which the TER exceeds $z$ in the stochastic SIR model with seasonal transmission, for a range of values of $M$ and $z$.** A. The TER (obtained by solving systems of equations (11) in the main text numerically) for a range of different values of the threshold number of infections, $M$. The blue shaded region shows the period of the year for which the TER exceeds $z = 0.4$ when $M = 100$. B. The duration of the year for which the TER exceeds $z = 0.4$, shown as a function of $M$. C. Heatmap indicating the duration of the year for which the TER exceeds $z$, shown for a range of values of $M$ and $z$. In all panels, values of $\beta_0 = 4$, $\beta_1 = 5$ and $\gamma = 4.9$ month$^{-1}$ are used. The overall population size was assumed to be $N = 1{,}000$ individuals.

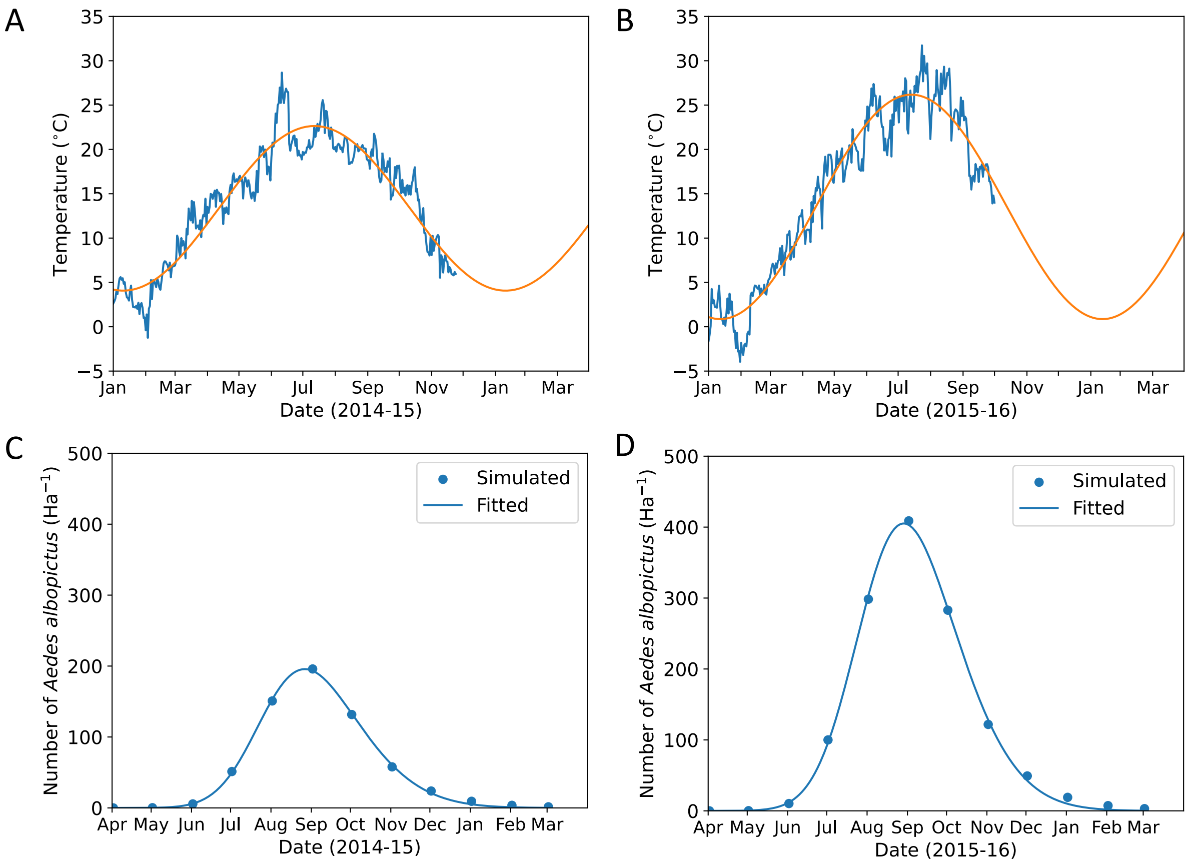


**Figure S1.4**. **Temperature and inferred vector density in Feltre, northern Italy, in 2014 and 2015.** A. Daily mean temperature in Feltre in 2014 (from MODIS satellite Land Surface Temperature measurements as described in [4]; blue line) and fitted temperature values obtained by fitting equation (4) in the main text to those data (orange line). B. Analogous to panel A, but using temperature data from 2015. C. Monthly number of adult female vectors per hectare in 2014 (and early 2015) obtained by solving system of equations (3) in the main text numerically based on the fitted temperature values in panel A (blue dots), and inferred number of adult female vectors per hectare obtained by fitting equation (5) in the main text to the monthly values (blue line). The ecological model is initialised at the beginning of April 2014, following the approach of Guzzetta *et al.* [4]. D. Analogous to panel C, but for 2015 (and early 2016), based on the fitted temperature values in panel B.
